# Repeat Hospitalizations and Their Costs in the 6 Months After Acute Myocarditis

**DOI:** 10.64898/2026.10.21.26366179

**Authors:** Ranjit Sah, Jaffar Hilli, Navya Pillikunte Doddareddy, George P Sorescu

## Abstract

**Background:** Readmission metrics after acute myocarditis usually record only whether the first readmission occurs within 30 days. The amount of inpatient care received after the first return and the period over which it was received are unknown.

**Methods:** We identified adults with a principal diagnosis of acute myocarditis (ICD-10-CM I40.x) in the Nationwide Readmission Database (NRD) from 2016 to 2022. The first qualifying admission in each patient year served as the index, and patients had to be discharged alive and not transferred. We linked every subsequent same-state, same-year admission for any cause within 30, 90, and 180 days. Using survey-weighted methods, we split the return length of stay (LOS) and estimated hospital cost (charges × cost-to-charge ratio, 2022 US dollars) into first-return and beyond-first-return components per index patient; patients without readmission were counted as having zero. The National Inpatient Sample (NIS) provided data on index mortality and costs.

**Results:** Of the 3,276 sampled index patients eligible for the 180-day follow-up (5,897 for 30 days), 6.5% were readmitted within 30 days and 14.0% within 180 days. Over 180 days, readmissions added 1.51 (95% CI, 1.23–1.79) bed-days and $5,884 (95% CI, 4,352–7,417) per index patient. Readmissions after the first contributed 0.62 (0.42–0.82) days and $2,430 (1,599–3,262), which is 41% of return bed-days and 41% of return costs (95% CI, 34–49 and 33–49, respectively). Counting only the first readmission, therefore, understated the 180-day return burden by 1.7-fold. The beyond-first share rose from 10% at 30 days to 32% at 90 days and 41% at 180 days, and 63% of the 180-day return bed-days accrued after day 30. Most readmissions had a non-cardiovascular (55% of bed-days) or other cardiovascular (33%) principal diagnosis, and myocarditis accounted for 3.5%. In the NIS (2,964 admissions; 71 in-hospital deaths), survivors had a mean index LOS of 4.4 days and a mean cost of $23,867.

**Conclusions:** After acute myocarditis, approximately two-fifths of the 6-month readmission bed-days and costs were from second and later readmissions. Most accrued after the usual 30-day window and were diagnoses other than myocarditis. First-readmission 30-day metrics substantially understate the post-discharge hospital burden in this population cohort.

**What Is Known:**

- Hospitalization for acute myocarditis carries a substantial risk of early readmission; national studies have reported 30- or 90-day readmission rates.
- Hospital quality and cost metrics typically consider only the first readmission within 30 days of discharge.

**What the Study Adds:** In a nationally representative US cohort, 41% of 180-day readmission bed-days and costs after acute myocarditis came from second and later readmissions; first-readmission metrics understated the 180- day return burden 1.7-fold.

Nearly two-thirds of the 180-day return bed-days occurred after day 30, and almost 9 in 10 were for diagnoses other than myocarditis, heart failure, or arrhythmia. These findings support follow-up that extends beyond 30 days and address coexisting conditions.

## Introduction

Acute myocarditis is an inflammatory disease of the myocardium. Presentation ranges from chest pain with preserved ventricular function to heart failure (HF), life-threatening ventricular arrhythmia, and cardiogenic shock.^1–3^ Most patients recover, but a meaningful proportion are rehospitalized after discharge.^3,4^ Analyses of US administrative data have described index outcomes, sex differences, and 30- or 90-day first readmissions after myocarditis.^4–9^

First-readmission endpoints indicate whether a patient returns, not how much hospital care that patient uses.^10,11^ A patient readmitted three times counts the same as one readmitted once, and admissions after day 30 are ignored.^12,13^ In conditions marked by recurrent instability or several coexisting diseases, these conventions can understate both the burden on patients and the resources a health system must plan for. Whether this is important after myocarditis, which mainly affects younger adults,^14,15^ has not been quantified.

We used the Nationwide Readmissions Database (NRD) to separate cumulative inpatient bed-days and hospital costs during the 30, 90, and 180 days after an index myocarditis admission, considering the contributions of the first and later readmissions, and to describe when and for which principal diagnoses this burden accrued. The National Inpatient Sample (NIS) provided a description of index hospitalization, including in-hospital mortality.

## Methods

### Data Availability

The HCUP data are available for purchase from the Agency for Healthcare Research and Quality (AHRQ) and cannot be shared by the authors under the HCUP Data Use Agreement. The complete Stata analysis code is available from corresponding author on reasonable request.

### Data Sources

The NRD is an all-payer database of inpatient discharges from participating states with verified patient linkage numbers that allow tracking of hospitalizations within a state and calendar year; weights produce national estimates.^16^ The NIS is a 20% stratified sample of US community hospital discharges.^17^ We used the NRD 2016–2022 and NIS 2016–2022 (with NIS 2023 as a descriptive extension). The two databases were analyzed separately and were not linked to each other.

### Study Population

We included adults aged ≥ 18 years with a principal ICD-10-CM diagnosis of acute myocarditis (I40.0, I40.1, I40.8, I40.9). In the NRD, the index admission was the first qualifying admission for each linked patient in each calendar year. We chose it before applying any outcome-based exclusion, so a later admission could not stand in for an ineligible first one. For follow-up analyses, we required a live discharge, a known non-transfer disposition, valid admission timing and LOS, and a non-combined (non-same-day-event) index record. To allow complete follow-up within the calendar year, we limited index discharges to January–November for 30-day analyses, January–September for 90- day analyses, and January–June for 180-day analyses.^18^ Pregnancy-associated admissions were retained.

### Outcomes

A readmission was defined as any later admission for any cause that began 1–30, 90, or 180 days after index discharge. For each index patient, we summed (1) the LOS and (2) the estimated cost of all readmissions, of the first readmission, and of readmissions after the first (“beyond-first”). Each readmission contributed its entire stay, assigned to the window in which it began (admission-anchored). Patients with complete follow-up and no readmission contributed zero; therefore, the means are per eligible index patient, not per readmitted patient. Cost was total charges multiplied by the hospital-specific HCUP cost-to-charge ratio,^19^ inflated to 2022 US dollars with the Bureau of Labor Statistics Consumer Price Index for Medical Care (series CUUR0000SAM, annual averages).^20^ Patients with a readmission that lacked valid charges, a cost-to-charge ratio, or clear hospital attribution were excluded from cost analyses only (complete-case domain); unknown values were never set to zero.

The principal diagnoses for readmission were classified as acute myocarditis, HF, ventricular arrhythmia, or complete atrioventricular block, or other cardiovascular, noncardiovascular, or invalid/unknown causes. In the 180-day cohort, we also split the return burden by the timing of readmission (days 1–30, 31–90, 91–180) within the same January–June cohort.

The secondary outcomes were any readmission and the number of readmissions (0, 1, or at least two) within each window. In the NIS, the outcomes were in-hospital mortality, LOS, and cost of the index admission, overall and by coded HF and electrical (ventricular tachycardia/fibrillation or complete heart block) phenotypes.

### Covariates

We described age, sex, race, ethnicity (NIS), primary payer, ZIP-code income quartile, patient residence urbanicity, the Charlson comorbidity index (Quan ICD-10 coding)^21,22^, and HF/electrical phenotype at the index.

### Statistical Analysis

All estimates used a complex survey design (hospital-year primary sampling units, database-year strata, and discharge weights) with Taylor-series linearization and subpopulation estimation over the full hospital roster.^23–25^ We estimated first-return, beyond-first, and all-return means jointly to preserve their covariance and then derived the beyond-first share (beyond-first/all-return) and all-to-first ratio using the delta method. Proportions were logit-transformed using 95% CIs.

We related the HF/electrical phenotype to 90-day readmission, 180-day return LOS, and 180-day return cost with survey-weighted logistic or linear regression adjusted for age, sex, Charlson index (excluding HF), payer, income, and urbanicity and year. The results are standardized (marginal) differences with Benjamini–Hochberg adjustment across the 9 prespecified contrasts.^26^ Because the NIS held only 71 index deaths, mortality models used a single parsimonious exploratory specification (age, sex, Charlson index, year), reported in the Supplement. Race- and payer-stratified mortality models had fewer than 10 events per parameter; we judged them underpowered and did not interpret them.

Sensitivity analyses included a broader definition, adding principal I51.4, restriction to I40.9, exclusion of concurrent acute myocardial infarction or COVID-19,^27^ exclusion of 2020, residents only, and alternative attribution of combined records (Table S5). Analyses were performed using Stata version 19 (StataCorp LLC, College Station, TX). In accordance with the HCUP Data Use Agreement, no statistic based on 10 or fewer unweighted observations was reported, with complementary suppression where needed. The study used de-identified data and was exempt from institutional review board review. Reporting follows the RECORD statement.^28^

## Results

### Study Population

The NIS contained 2,964 sampled adult admissions with a principal diagnosis of acute myocarditis and known vital status (2,893 survivors and 71 in-hospital deaths; weighted in-hospital mortality, 2.4% [95% CI, 1.9–3.0]). In the NRD, 5,897, 4,783, and 3,276 sampled index patients were eligible for the 30-, 90-, and 180-day follow-up (Figure S1), corresponding to approximately 1,540, 1,250, and 850 index hospitalizations per year, respectively. The return LOS was complete for all 3,276 patients in the 180-day cohort, and the return cost was complete for 3,248 (99.1%) patients. Table 1 shows the index characteristics: mean age, 41.1 years; 65% male; and 30% with HF codes.

**Table 1.** Characteristics of Adults Hospitalized With Acute Myocarditis, NIS 2016–2022.

| Characteristic | Overall | Survived | Died in hospital |
| --- | --- | --- | --- |
| Unweighted sampled admissions, n | 2,964 | 2,893 | 71 |
| Age, y, mean (95% CI) | 41.1 (40.5–41.8) | 40.8 (40.1–41.5) | 55.0 (50.4–59.6) |
| **Age group, %** |  |  |  |
| 18–34 | 45.8 (44.0–47.6) | 46.4 (44.5–48.3) | 21.1 (13.3–31.9) |
| 35–44 | 15.8 (14.5–17.2) | 16.0 (14.7–17.4) | Suppressed* |
| 45–54 | 13.3 (12.1–14.6) | 13.2 (12.0–14.5) | 18.3 (11.0–28.9) |
| 55–64 | 11.3 (10.2–12.5) | 11.2 (10.1–12.4) | Suppressed* |
| 65 or older | 13.8 (12.6–15.1) | 13.2 (12.0–14.5) | 39.4 (28.7–51.3) |
| Male, % | 64.7 (63.0–66.5) | 65.0 (63.2–66.7) | 53.5 (42.1–64.6) |
| **Race and ethnicity, %** |  |  |  |
| White / Black / Hispanic | 60.3 / 14.8 / 13.5 | 60.4 / 14.7 / 13.8 | 57.7 / 22.5 / Supp.* |
| Asian or Pacific Islander / Native American / Other / Unknown | 3.2 / 0.5 / 3.8 / 3.9 | 3.0 / 0.5 / 3.9 / 3.8 | Supp.* / 0 / Supp.* / Supp.* |
| **Primary payer, %** |  |  |  |
| Medicare / Medicaid / Private | 14.4 / 17.6 / 55.2 | 13.9 / 17.7 / 55.6 | 35.2 / 15.5 / 38.0 |
| Self-pay / No charge / Other / Unknown | 8.3 / Supp.* / 3.7 / Supp.* | 8.3 / Supp.* / 3.7 / Supp.* | Supp.* / 0 / Supp.* / 0 |
| ZIP income quartile 1 (lowest) to 4, % | 23.4 / 23.5 / 24.8 / 26.9 | 23.5 / 23.6 / 24.7 / 26.8 | 22.5 / Supp.* / 28.2 / 28.2 |
| Charlson index, mean (95% CI) | 1.22 (1.16–1.28) | 1.17 (1.11–1.23) | 3.15 (2.50–3.81) |
| **Coded cardiac phenotype at index, %** |  |  |  |
| Neither HF nor electrical syndrome | 66.7 (64.9–68.4) | 67.7 (66.0–69.4) | Suppressed* |
| HF only | 23.0 (21.5–24.6) | 22.8 (21.3–24.4) | 32.4 (22.7–43.9) |
| Electrical syndrome only | 3.7 (3.1–4.5) | 3.6 (3.0–4.4) | Suppressed* |
| HF and electrical syndrome | 6.5 (5.7–7.5) | 5.8 (5.0–6.8) | 35.2 (25.2–46.7) |
| **Index resource use, mean (95% CI)** |  |  |  |
| Length of stay, d | 4.66 (4.29–5.03) | 4.38 (4.07–4.69) | 16.08 (7.59–24.58) |
| Hospital cost, 2022 US\$ | 27,190 (23,869–30,511) | 23,867 (21,327–26,406) | 165,483 (78,111–252,855) |
| Charges, 2022 US\$ | 113,605 (98,806–128,404) | 101,074 (88,314–113,834) | 634,988 (325,164–944,813) |

### Index Hospitalization

The mean index LOS was 4.38 days (95% CI, 4.07–4.69) among survivors and 16.08 days (7.59– 24.58) among patients who died. The mean index cost among survivors was $23,867 (21,327– 26,406). The index LOS increased with coded cardiac complications: 2.59 days (2.36–2.83) days with neither HF nor an electrical syndrome, 6.75 days (6.04–7.47) days with HF only, and 15.44 days (12.89–18.00) days with both (Table S1).

### Readmission Occurrence and Multiplicity

Within 30, 90, and 180 days, 6.5%, 10.5%, and 14.0% of the index patients were readmitted, and 4.7% had two or more readmissions within 180 days (Table 2). Among patients readmitted within 180 days, the mean cumulative return LOS was 10.8 (95% CI, 9.1–12.6) days and the mean return cost was $44,260 (95% CI, 33,435–55,085).

**Table 2.** Readmission Occurrence and Return Resource Burden per Index Patient, NRD 2016–2022.

| **Measure** | **30 days** | **90 days** | **180 days** |
| --- | --- | --- | --- |
| Eligible sampled index patients, n | 5,897 | 4,783 | 3,276 |
| **Readmission occurrence** |  |  |  |
| Any readmission, % (95% CI) | 6.5 (5.8–7.2) | 10.5 (9.6–11.4) | 14.0 (12.7–15.3) |
| 2 or more readmissions, % (95% CI) | 0.7 (0.5–1.0) | 2.7 (2.2–3.2) | 4.7 (4.0–5.6) |
| **Return length of stay, days per index patient** |  |  |  |
| All returns | 0.47 (0.39–0.55) | 1.00 (0.82–1.18) | 1.51 (1.23–1.79) |
| First return | 0.42 (0.35–0.50) | 0.68 (0.58–0.79) | 0.89 (0.74–1.04) |
| Returns after the first | 0.05 (0.03–0.07) | 0.32 (0.19–0.44) | 0.62 (0.42–0.82) |
| Beyond-first share, % (95% CI) | 10 (6–14) | 32 (23–40) | 41 (34–49) |
| **Return hospital cost, 2022 US\$ per index patient** | | | |
| All returns | 1,914 (1,380–2,447) | 3,808 (2,906–4,710) | 5,884 (4,352–7,417) |
| First return | 1,787 (1,261–2,312) | 2,749 (2,052–3,446) | 3,454 (2,496–4,411) |
| Returns after the first | 127 (67–186) | 1,059 (627–1,491) | 2,430 (1,599–3,262) |
| Beyond-first share, % (95% CI) | 7 (3–10) | 28 (19–36) | 41 (33–49) |
| **Index plus returns, 180-day cohort** |  |  |  |
| Total inpatient LOS, d | – | – | 5.22 (4.84–5.59) |

### Burden Beyond the First Readmission

Over 180 days, readmissions added a mean of 1.51 bed-days (95% CI, 1.23–1.79) per index patient: 0.89 (0.74–1.04) from first readmissions and 0.62 (0.42–0.82) from later readmissions (Table 2; Figure 1A). Readmissions after the first thus made up 41% (95% CI, 34–49) of 180-day return bed-days, and all-return LOS was 1.70 times (95% CI, 1.48–1.91) the first-return LOS. Costs followed the same pattern: $5,884 (4,352–7,417) for all returns, $3,454 (2,496–4,411) for first returns, and $2,430 (1,599–3,262) for returns after the first or 41% of return costs (Figure 2). With the index stay included, the average total 180-day inpatient LOS was 5.22 days (4.84–5.59), with readmissions contributing 29%.

**Figure 1.**
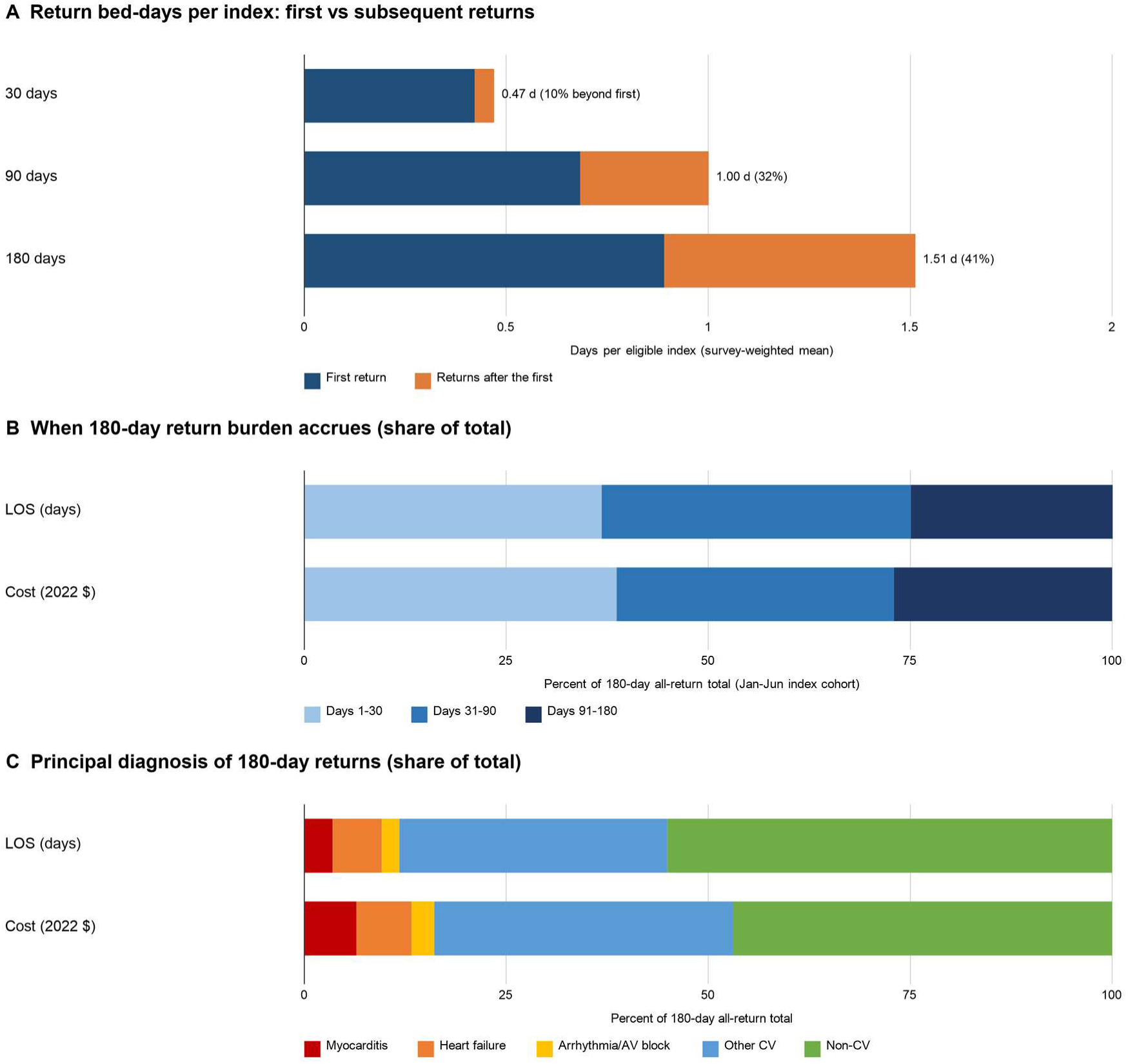
Decomposition of inpatient readmission burden after acute myocarditis. **A**, Mean return bed-days per eligible index patient from the first readmission and from subsequent readmissions within 30, 90, and 180 days (separately eligible cohorts); labels give the total and beyond-first share. **B**, Share of 180-day return bed-days and costs accruing in days 1–30, 31–90, and 91–180 after discharge. **C**, Share of 180-day return bed-days and costs by principal diagnosis of the readmission. NRD 2016–2022; survey-weighted estimates; 95% CIs in Tables 2 and 3. AV, atrioventricular; CV, cardiovascular.

**Figure 2.**
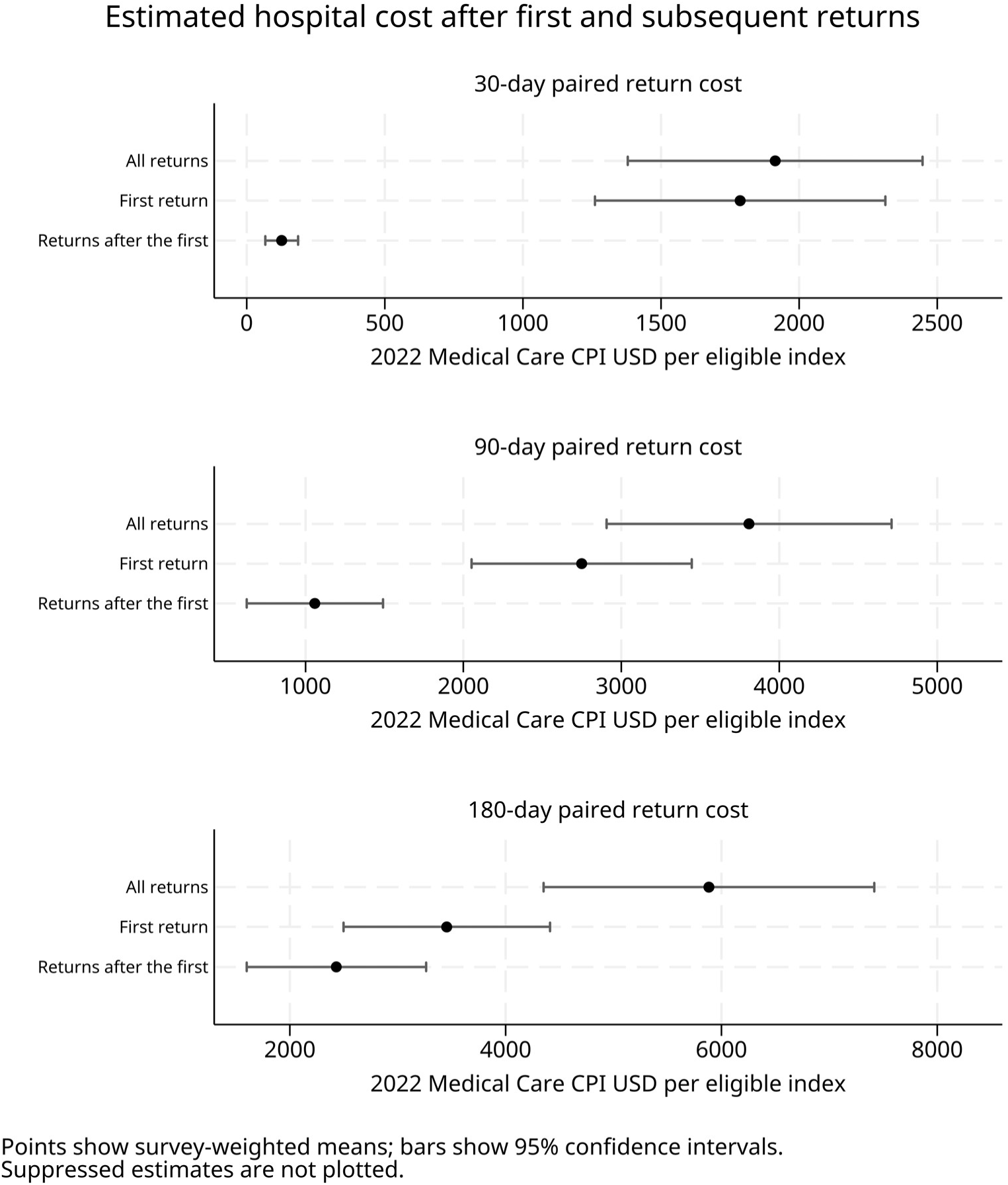
Hospital cost of first and subsequent readmissions per eligible index patient within 30, 90, and 180 days. Survey-weighted means with 95% CIs, 2022 US dollars (Medical Care CPI). Patients without readmission contribute zero.

**Table 3.** 180-Day Return Burden by Principal Diagnosis and Timing of Readmission.

| Principal diagnosis of readmission | Days 1–30 | Days 31–90 | Days 91–180 | Total (share) |
| --- | --- | --- | --- | --- |
| **Length of stay, days per index patient** |  |  |  |  |
| Acute myocarditis | 0.041 (0.003–0.079) | 0.011 (-0.009–0.030) | 0.001 (-0.001–0.003) | 0.05 (3.5%) |
| Heart failure | 0.039 (0.010–0.068) | 0.039 (-0.004–0.081) | 0.014 (0.004–0.025) | 0.09 (6.1%) |
| Ventricular arrhythmia / complete AV block | 0.011 (0.001–0.020) | 0.011 (-0.004–0.025) | 0.012 (0.000–0.023) | 0.03 (2.2%) |
| Other cardiovascular | 0.209 (0.115–0.303) | 0.197 (0.117–0.277) | 0.096 (0.064–0.129) | 0.50 (33.2%) |
| Noncardiovascular | 0.257 (0.188–0.327) | 0.322 (0.157–0.486) | 0.254 (0.175–0.333) | 0.83 (55.1%) |
| All diagnosis groups* | 0.56 (0.43–0.68) | 0.58 (0.38–0.77) | 0.38 (0.29–0.47) | 1.51 (100%) |
| **Hospital cost, 2022 US\$ per index patient** | | | | |
| Acute myocarditis | 268 (-58–595) | 108 (-101–316) | 4 (-3–11) | 380 (6.5%) |
| Heart failure | 308 (-50–666) | 33 (9–57) | 60 (13–108) | 401 (6.8%) |
| Ventricular arrhythmia / complete AV block | 53 (-9–116) | 40 (-25–104) | 73 (-13–158) | 166 (2.8%) |
| Other cardiovascular | 851 (238–1,464) | 937 (333–1,540) | 390 (236–543) | 2,177 (37.0%) |
| Noncardiovascular | 794 (559–1,030) | 903 (455–1,351) | 1,063 (561–1,565) | 2,760 (46.9%) |
| All diagnosis groups* | 2,275 | 2,020 | 1,589 | 5,884 (100%) |

January–June index cohort eligible for 180-day follow-up. Survey-weighted means (95% CI). Lower confidence limits below zero are Wald limits around near-zero means. \*Includes invalid/unknown principal diagnosis, not shown separately because of HCUP cell-size rules; segment totals for cost are sums of displayed components.

The beyond-first contribution grew with the length of the window: 0.05 days (0.03–0.07) at 30 days, 0.32 (0.19–0.44) at 90 days, and 0.62 (0.42–0.82) at 180 days, or 10%, 32%, and 41% of return bed-days, respectively (Figure 1A). Each window had its own discharge-month eligibility; therefore, these comparisons were made across cohorts, not within one cohort.

### Timing and Principal Diagnoses of Return Burden

In the 180-day cohort, return bed-days per index patient were 0.56 (0.43–0.68) in days 1–30, 0.58 (0.38–0.77) in days 31–90, and 0.38 (0.29–0.47) in days 91–180. After day 30, 63% of bed-days and 61% of costs were incurred (Figure 1B; Table 3).

Noncardiovascular principal diagnoses accounted for 0.83 (0.62–1.04) return bed-days per index patient (55%), other cardiovascular diagnoses for 0.50 (0.37–0.63; 33%), HF for 0.09 (6%), and ventricular arrhythmia or complete heart block for 0.03 (2%). Readmissions coded as acute myocarditis contributed 0.05 days (0.01–0.10; 3.5%), mostly in the first 30 days (Figure 1C; Table 3). Beyond-first bed-days were also mainly noncardiovascular (0.36 [0.19–0.54]) or other cardiovascular (0.20 [0.12–0.27]) complications. Costs were distributed similarly (noncardiovascular 47%, other cardiovascular 37%, HF 7%, and myocarditis 6%).

### Phenotype Associations and Sensitivity Analyses

Compared with patients with neither coded HF nor an electrical syndrome, patients with HF only had 7.2 percentage points (95% CI, 4.7–9.7) higher 90-day readmission, 1.98 (95% CI, 0.69–3.26) more 180-day return bed-days, and $10,402 (95% CI, 3,453 to 17,351) higher 180-day return cost (Benjamini-Hochberg adjusted P<0.001, P=0.006, and P=0.006). Patients with both HF and an electrical syndrome had corresponding differences of 18.0 percentage points (11.3 to 24.6), 4.52 days (1.80 to 7.23), and $26,826 (5,355 to 48,298) (adjusted P<0.001, P=0.003, and P=0.02). Electrical syndrome alone was not associated with any of the three outcomes (-1.2 percentage points [-4.6 to 2.3], 0.19 days [-0.84 to 1.23], and $44 [-2,852 to 2,941]; adjusted P=0.65, P=0.80, and P=0.98) (Table S3). In sensitivity analyses, the 180-day all-return LOS per index patient was 1.69 (1.00–2.37) days with the I40.9-only definition, 1.49 (1.21–1.77) days after excluding concurrent acute myocardial infarction, and 1.46 (1.28–1.64) days with the broader definition including I51.4; the beyond-first share was 47%, 42%, and 42%, respectively, and the direction of every finding remained unchanged (Table S5).

## Discussion

In this national cohort of adults hospitalized with acute myocarditis, second and later readmissions generated approximately two-fifths of all inpatient bed-days and hospital costs incurred during the 6 months after discharge. Counting only the first readmission understated the 180-day return burden 1.7-fold, and a 30-day window captured little more than one-third of it. Most of this burden was due to principal diagnoses other than myocarditis.

These findings extend the national studies of myocarditis readmission that reported first readmissions within 30 or 90 days.^4–9^ Such metrics suit quality benchmarking but answer a narrower question than ours. A single return does not capture the course after discharge: the beyond-first share rose from 10% at 30 days to 41% at 180 days, indicating that a subset of patients was readmitted repeatedly over months. Payers and health systems estimating the consequences of myocarditis, and investigators designing outcome studies, should consider cumulative and longer endpoints, such as total hospital days or recurrent-event analyses, rather than time to first readmission alone.^10,11,29,30^

The diagnoses behind these returns were clinically significant. Readmissions coded principally as myocarditis contributed only 3.5% of bed-days, and HF and arrhythmia together contributed under 10%. Noncardiovascular and other cardiovascular diagnoses dominated, including among later returns.^31,32^ Administrative codes cannot separate sequelae of myocarditis from unrelated illnesses, and some cardiovascular returns (pericarditis or chest pain evaluations, for example) may relate to the index condition. However, the data suggest that post-discharge care, in line with current guidelines emphasizing structured follow-up,^1,33^ should address coexisting conditions and the cause of myocarditis (systemic inflammatory, infectious, or drug-related disease, for instance),^2,34^ not only ventricular function and rhythm.

Because 63% of 180-day return bed-days accrued after day 30, interventions and quality metrics tied to the 30-day window may miss most of the opportunities.^13^ These data cannot show whether any of this burden is preventable or not. Studies linking inpatient records with outpatient care, imaging, and the cause of myocarditis are needed to identify patients at risk of repeated returns.

### Limitations

This study has limitations. First, myocarditis was identified from principal discharge codes without imaging or biopsy confirmation. Validation studies suggest a moderate positive predictive value,^35,36^ and the results were similar with a restricted I40.9 definition and after excluding concurrent myocardial infarction (beyond-first share 47% and 42%, respectively; Table S5). Second, the NRD follows readmissions only within the same state and calendar year. Readmissions in another state, in the following year, or with discharge after December 31 were missed, as were observation stays, emergency visits, outpatient care, and out-of-hospital deaths. To ensure complete follow-up, we restricted index discharges by month, which excludes later-year index admissions and may introduce seasonal selection. Third, costs came from charges and hospital-level cost-to-charge ratios; they represent hospital production costs, not payments or societal costs. Fourth, cost analyses excluded patients with incomplete charge or attribution data (0.9% at 180 days), and this missingness may not be random. Fifth, NRD estimates count patient-years rather than unique individuals because linkage resets each calendar year. Sixth, the small number of in-hospital deaths precluded reliable adjusted analyses of mortality disparities. Finally, survey-based Wald intervals for small components can extend below zero; this reflects uncertainty around near-zero means.

## Conclusions

After hospitalization for acute myocarditis, readmissions beyond the first account for approximately 41% of 6-month readmission bed-days and costs. Most of the return burden accrued after 30 days, and most were for diagnoses other than myocarditis. First-readmission and 30-day metrics substantially understate the post-discharge hospital burden of myocarditis.

## Acknowledgments

The authors acknowledge the HCUP Data Partners who contributed to the NIS and NRD.

## Supplemental Material

Tables S1–S6, Figure S1, RECORD checklist and analysis code.

## AI Declaration

Paperpal, ChatGPT and Claude were used only for language and stylistic refinement. These tools were not involved in study design, data analysis, interpretation, or content development. All intellectual contributions and scientific interpretations are the sole work of the authors, who take full responsibility for the accuracy, originality, and integrity of the manuscript.

## Authors Contributions

RS and GPS developed the idea, do the analysis, and purposed the initial draft, NPD, and JH critical edit the manuscript. All authors read and approve for the final manuscript.

## Supplemental Material

Table S1. Index LOS, charges, and costs by HF/electrical phenotype and vital status (NIS), including NIS 2023 extension.

Table S2. Charlson and Elixhauser comorbidity prevalence (NIS and NRD).

Table S3. Standardized differences by index HF/electrical phenotype in 90-day readmission and 180-day return LOS and cost, with Benjamini–Hochberg adjusted P values (9 prespecified contrasts).

Table S4. Return resources in nominal dollars and charges; index-plus-return totals; conditional means among readmitted patients.

Table S5. Sensitivity analyses: broad definition (I51.4), I40.9 only, excluding acute MI, excluding COVID-19, excluding 2020, residents only, first-documented-adult ascertainment, strict/last-hospital cost attribution, strict month cutoffs.

Table S6. Exploratory in-hospital mortality models (NIS), with events-per-parameter reported.

Figure S1. Cohort derivation flow diagram (NIS and NRD), with exclusions at each step (all cells >10 or suppressed).

Figure S2. Return LOS and cost by principal diagnosis and readmission timing with 95% CIs (current Figure 3).

